# A new method for removing artifacts from recordings of the electrically evoked compound action potential: Single-pulse stimulation

**DOI:** 10.1101/2024.01.17.24301435

**Authors:** Jeffrey Skidmore, Yi Yuan, Shuman He

## Abstract

This report presents a new method for removing electrical artifact contamination from the electrically evoked compound action potential (eCAP) evoked by single cathodic-leading, biphasic-pulse stimulation. The development of the new method is motivated by results recorded in human cochlear implant (CI) users showing that the fundamental assumption of the classic forward masking artifact rejection technique is violated in up to 45% of cases tested at high stimulation levels when using default stimulation parameters. Subsequently, the new method developed based on the discovery that a hyperbola best characterizes the artifacts created during stimulation and recording is described. The eCAP waveforms obtained using the new method are compared to those recorded using the classic forward masking technique. The results show that eCAP waveforms obtained using both methods are comparable when the fundamental assumption of the classic forward masking technique is met. In contrast, eCAP amplitudes obtained using the two methods are significantly different when the fundamental assumption of the classic forward masking technique is violated, with greater differences in the eCAP amplitude for greater assumption violations. The new method also has excellent test-retest reliability (Intraclass correlation > 0.98). Overall, the new method is a viable alternative to the classic forward masking technique for obtaining artifact-free eCAPs evoked by single-pulse stimulation in CI users.

## INTRODUCTION

The electrically evoked compound action potential (eCAP) measured at the auditory nerve is a summed response generated by a group of auditory nerve fibers (ANFs) responding synchronously to electrical stimulation [1, 2]. This near-field response can be recorded directly from a patient’s cochlear implant (CI) using the telemetry functions implemented in the CI and the commercial software provided by the CI manufacturer. The eCAP has been shown to be useful for estimating the physiological status of the auditory nerve [3–9]. These estimates of the physiological status of the auditory nerve may have clinical benefits such as longitudinal monitoring of neural health [10, 11], implant fitting [12–14] and explaining variance in speech perception performance among CI users [9, 15–18].

The primary challenge in recording eCAPs is the presence of unwanted voltages (i.e., electrical artifacts) that contaminate and obscure the neural response. The largest artifact is caused by decaying charges produced during stimulation (i.e., stimulation artifact) due to capacitors in the CI [19] and the capacitive properties of the electrode– electrolyte interface [20]. The stimulation artifact increases with stimulation level and is typically several orders of magnitude larger than the eCAP. Another artifact comes from the switching of the recording amplifier during the measurement process (i.e., recording artifact). While smaller than the stimulation artifact, the recording artifact is sufficiently large that it can contaminate the eCAP response, especially at stimulation levels near the eCAP threshold. Therefore, techniques to remove or reduce the stimulation and recording artifacts from eCAP recordings are necessary.

Several artifact rejection techniques have been used or proposed over the past few decades for recording eCAPs in response to single-pulse stimulation. These techniques include the classic two-pulse forward masking [FwdMsk; 1], alternating polarity, and subthreshold template subtraction [21], along with more recent techniques such as precision triphasic-pulse stimulation [22], independent component analysis [23], and multi-curve-fitting [24]. The strengths, weaknesses, and limitations of each technique have been described previously [e.g., 2, 23, 25, 26].

In recent years, FwdMsk has by far been the most commonly used artifact rejection technique for eCAP recordings, especially in CI users [e.g., 3-5, 7-9, 12, 15, 27, 28-32]. However, while the important considerations and limitations of FwdMsk are well known in theory, it is difficult to choose appropriate stimulation parameters in practice because of the challenges in verifying the underlying assumptions of this technique when collecting eCAP data. Therefore, as the motivation for the development of the method described in this report, we first review the theoretical basis and demonstrate the limitations of FwdMsk before describing the new method.

## TWO-PULSE FORWARD MASKING

The classic two-pulse forward masking technique [1] has been used in many studies over the last few decades for recording eCAPs in CI users. The method creates templates of the stimulation and recording artifacts by recording voltages in response to four stimuli. The first stimulus is a single pulse (i.e., probe pulse) which results in a recorded voltage trace (‘A’ trace) that includes the probe stimulation artifact, the recording artifact, and the eCAP evoked by the probe. The second stimulus is the same as the first stimulus with the addition of a masker pulse which precedes the probe pulse by a specified inter-stimulus interval (i.e., masker-probe interval, MPI). In addition to the masker stimulation artifact and the neural response evoked by the masker pulse, the recorded voltage trace (‘B’ trace) consists of the probe stimulation artifact, the recording artifact, and potentially an eCAP response evoked by the probe pulse. Ideally, a relatively high masker stimulation level compared to the probe stimulation level and a short MPI are used to set the neurons in an absolute refractory state so that there is no neural response to the probe pulse. This is the fundamental assumption of FwdMsk. The third stimulus is the same as the second stimulus without a probe pulse. This single (masker) pulse results in a recorded voltage trace (‘C’ trace) that contains the masker stimulation artifact, the recording artifact, and the neural response evoked by the masker pulse. The fourth stimulus is a zero-amplitude stimulation that provides a template of the recording artifact (‘D’ trace) caused by the switching of the recording amplifier. After the four traces are recorded, a template waveform (‘T’ waveform) that consists of the probe stimulation artifact, the recording artifact, and any eCAP response evoked by the probe pulse in the second recording, is derived by adding the fourth recording to the difference between the second and third recordings (i.e., ‘T’ = ‘B’ – ‘C’ + ‘D’). Finally, the eCAP waveform (‘E’ waveform) is obtained by subtracting this artifact template from the first recording (i.e., ‘E’ = ‘A’ – ‘T’). Therefore, any neural response to the probe pulse included in the ‘B’ trace is also in the artifact template ‘T’ and alters the derived eCAP waveform ‘E’. This method is illustrated in Figure 1 for supra-threshold stimulation levels. Examples of each recorded trace and derived waveform are shown in Figure 2 for one case in which the artifact template is free of neural response (i.e., complete masking) and one case in which the artifact template has a neural response (i.e., incomplete masking).

**Figure 1.**
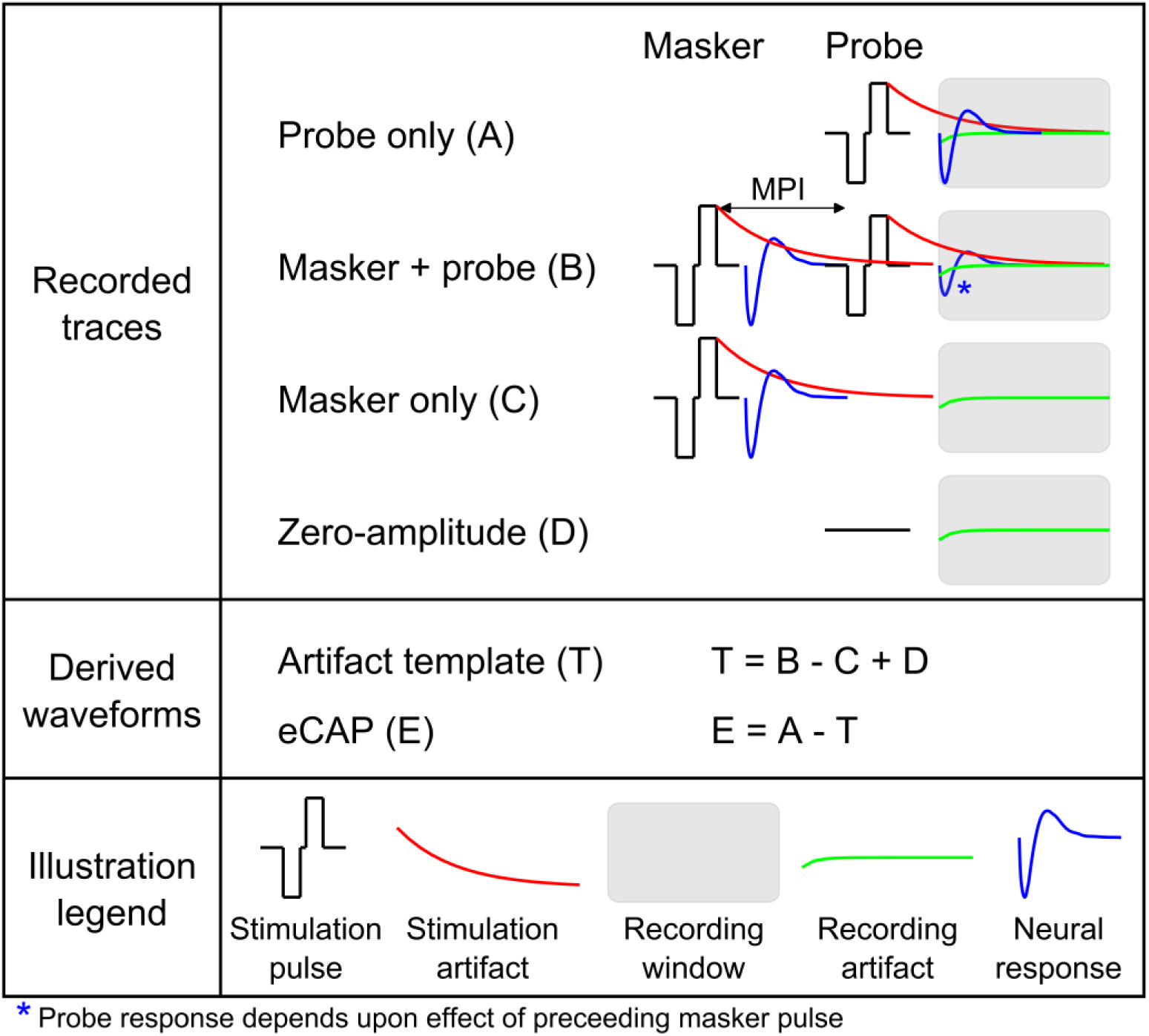
Illustration of the classic two-pulse forward masking artifact rejection technique for removing artifacts from recordings of the electrically evoked compound action potential (eCAP).

**Figure 2.**
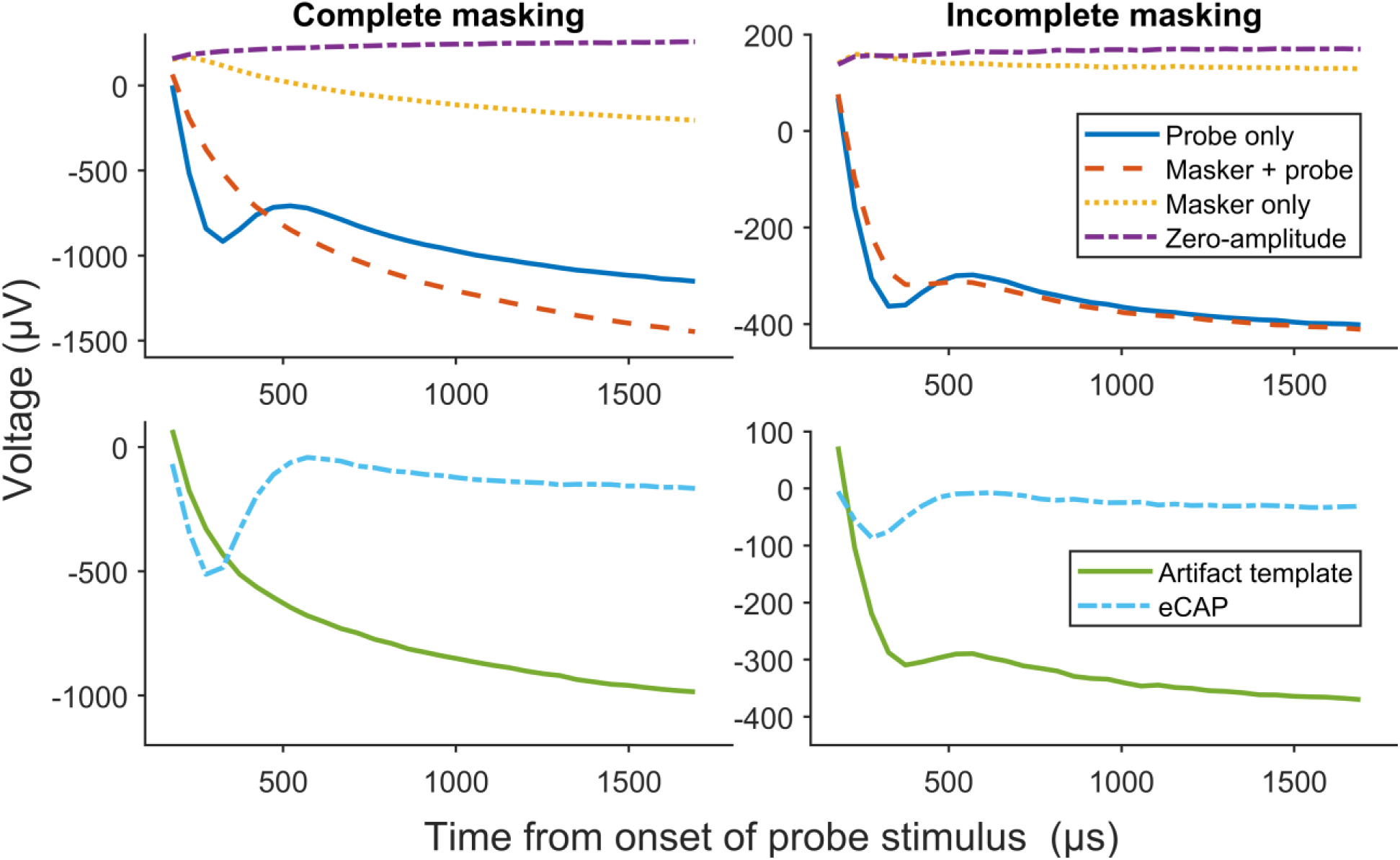
Examples of recorded traces and derived waveforms obtained using the classic two-pulse forward masking artifact rejection technique for one case in which the artifact template is free of neural response (left panel) and one case in which the artifact template has a neural response (right panel).

As stated above, FwdMsk only produces an artifact-free eCAP waveform if there is no neural response to the probe pulse when proceeded by a masking pulse. The two primary factors that affect the validity of this assumption are the stimulation level of the masker pulse relative to the stimulation level of the probe pulse and the duration of the MPI. The considerations and implications of each of these two factors are reported below.

### Stimulation Level of Masker Pulse

In FwdMsk, the masker pulse must be sufficiently large to activate the target ANFs. Otherwise, the neurons that are not activated by the masker pulse may be activated by the probe pulse that follows. This incomplete masking is manifest by neural response being present in the ‘B’ trace, as observed in the right panel of Figure 2, and results in a reduced eCAP compared to the fully-masked condition. A difference in stimulation level between the masker pulse and the probe pulse (i.e., masker offset) of +10 current levels (CL) has been proposed to be sufficient for producing the desired masking effect [33] and is a frequently used masker offset [e.g., 4, 6, 8, 9, 12, 27-30].

### Duration of Masker-probe Interval

In FwdMsk, the MPI must be equal or shorter than the absolute refractory period (ARP) of the target neurons so that all neurons activated by the masker pulse will not respond to the probe pulse. Otherwise, some neurons activated by the masker pulse may have recovered sufficiently to respond to the probe pulse and generate an action potential. The fraction of neurons that respond to the probe pulse is influenced by at least two factors: the difference between the MPI and the ARP, and the speed of recovery during the relative refractory period (RRP). More neurons respond to the probe pulse when there are larger differences between the MPI and the ARP and when there is faster recovery during the RRP. Therefore, there are larger neural responses in the artifact template with larger differences between the MPI and the ARP, as shown in the middle panel of Figure 3. Additionally, phase-shifts in the artifact templates relative to the probe-only recordings, likely due to cross-fiber variability in refractory recovery times [34], change the morphology of the derived eCAP waveforms. As observed in the bottom panel of Figure 3, there are increasing changes in peak latencies and peak amplitudes relative to the fully-masked condition (i.e., MPI = 300 µs) with increasing time between the MPI and the ARP. Therefore, eCAP amplitudes and peak latencies obtained using FwdMsk are not accurate if the MPI is longer than the ARP, with larger errors occurring for larger differences between the MPI and the ARP.

**Figure 3.**
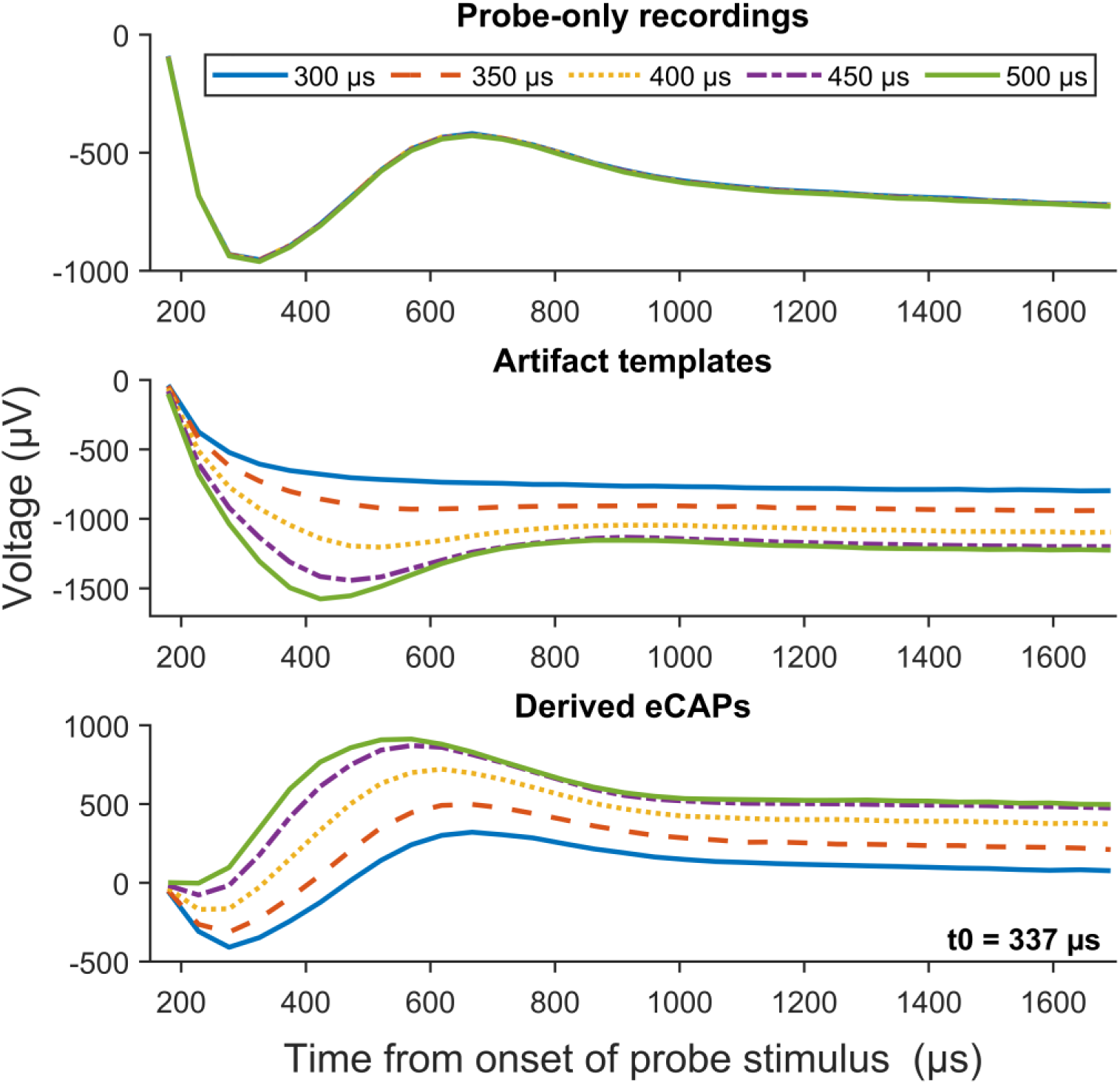
Recorded traces to probe-only stimulation (top panel) and derived artifact templates and eCAP waveforms (middle and bottom panel, respectively) obtained using the classic two-pulse forward masking artifact rejection technique for single-pulse stimuli with various masker-probe intervals at one electrode location in one adult cochlear implant user. The absolute refractory recovery period estimated at this electrode location (i.e., t0) is provided in the corner of the bottom panel.

In practice, it is not straightforward to verify whether the MPI is set correctly when collecting eCAP data in individual CI users because it requires an estimate of the ARP. Therefore, it is not well-known how often the assumption of complete masking is met when using FwdMsk. Moreover, recovery from refractoriness is affected by the stimulation level, with faster recovery occurring at higher stimulation levels [34, 35]. At low stimulation levels and short MPIs (i.e., < 300 µs), a faciliatory effect could occur in which neurons not activated by the first pulse could be activated by the second pulse due to temporal integration of the charge [36]. The strongest facilitation effect is observed when the first pulse is near the eCAP threshold [35, 37], and the effect increases for shorter MPIs [38]. Refractory recovery periods and facilitatory effects are also influenced by the health of the ANFs [39, 40]. Therefore, the optimal MPI for recording eCAPs using FwdMsk could vary across CI users, electrode locations and stimulation levels.

### Absolute Refractory Periods at High Stimulation Levels

As discussed previously, FwdMsk is dependent on the MPI being within the ARP of the target ANFs. An MPI of 400 µs has been used frequently in eCAP studies in CI users with Cochlear™ Nucleus® or Advanced Bionics devices [e.g., 3-5, 7-9, 12, 15, 25, 27, 28-32, 40]. Therefore, it is important to understand whether 400 µs is generally within the ARP, and therefore, an appropriate MPI for recording eCAPs in CI users. The ARP (i.e., the time period following stimulation in which none of the target neurons could generate an action potential) can be estimated by fitting an exponential decay function to the eCAP refractory recovery function as has been done in previously published studies [e.g., 8, 9, 19, 40, 41]. The eCAP refractory recovery function is obtained using the modified template subtraction technique [26]. The modified template subtraction technique is a modification of FwdMsk that enables the measurement of artifact-free eCAPs obtained in a paired-pulse stimulation paradigm with various MPIs. Importantly, the default reference MPI used in the modified template subtraction technique is 300 µs, instead of 400 µs used in FwdMsk. Using the modified template subtraction technique, Morsnowski, et al. (41) reported a median ARP of 390 µs across 84 electrode locations measured in 14 CI users when evaluated at the participant’s maximum comfort level (i.e., C level). Therefore, more than half of the electrode locations in their study had an ARP of less than 400 µs when measured at C level.

To confirm the finding reported in Morsnowski, et al. (41), we evaluated ARPs at 473 electrode locations across 80 pediatric and adult CI users (Pediatrics: 27 participants, 38 ears, 127 electrode locations; Adults: 53 participants, 62 ears, 346 electrode locations). All participants used a Cochlear™ Nucleus® device (Cochlear Ltd.) and had normal inner ear anatomy. eCAP recordings were obtained using the Advanced Neural Response Telemetry function implemented in the Custom Sound EP (v. 5.1, v.5.2 or v.6.0) commercial software (Cochlear Ltd, Sydney, NSW, Australia) using the modified template subtraction technique [26]. Both the masker and the probe were symmetric, cathodic-leading, biphasic pulses with an interphase gap of 7 µs and a pulse phase duration of 25 µs/phase. The masker and the probe were presented to the test electrode at the participants’ C level and 10 CL below C level, respectively, to obtain a masker offset of +10 CL. eCAPs were recorded as the MPI was systematically increased from 100 μs to 10 ms. Other recording parameters included a 122-µs recording delay, an amplifier gain of 50 dB, and a sampling rate of 20,492 Hz. Estimates of the ARP (i.e., t_0_) were obtained by fitting a decaying exponential function to the eCAP amplitudes plotted as a function of MPI as done in our previous studies [8, 9, 40]. Any estimates of the ARP below 300 µs were excluded as poor fits, as was done by Morsnowski, et al. (41), because of the use of 300 µs as the reference MPI in the modified template subtraction technique. In total, 16/473 (3.4%) of the estimates were excluded (Pediatrics: 3/127 = 2.4%; Adults 13/346 = 3.8%).

The ARP estimates measured in pediatric and adult CI users are shown separately in Figure 4. The result of a Mann-Whitney U test showed that estimated ARPs were significantly longer in the pediatric CI users than in the adult CI users (U = 17422, p = 0.010). This can be explained, at least in part, by the difference in stimulation levels used in these two participant groups. Specifically, the result of a Mann-Whitney U test showed that the stimulation level was significantly lower in the pediatric CI users than in the adult CI users (U = 16088, p < 0.001). There was a significant negative correlation between estimated ARP and stimulation level for both patient populations (Pediatrics: N = 124, r = −0.28, p = 0.002; Adults: N = 333, r = −0.31, p < 0.001), indicating shorter ARPs at higher stimulation levels. This is consistent with the results of other studies [34, 35]. More importantly, 192/457 (42.0%) of the estimated ARPs were less than 400 µs (Pediatrics: 41/124 = 33.1%; Adults: 151/333 = 45.4%). These data clearly indicate that FwdMsk is not sufficient for removing artifacts from eCAP recordings in many cases due to violated underlying assumptions.

**Figure 4.**
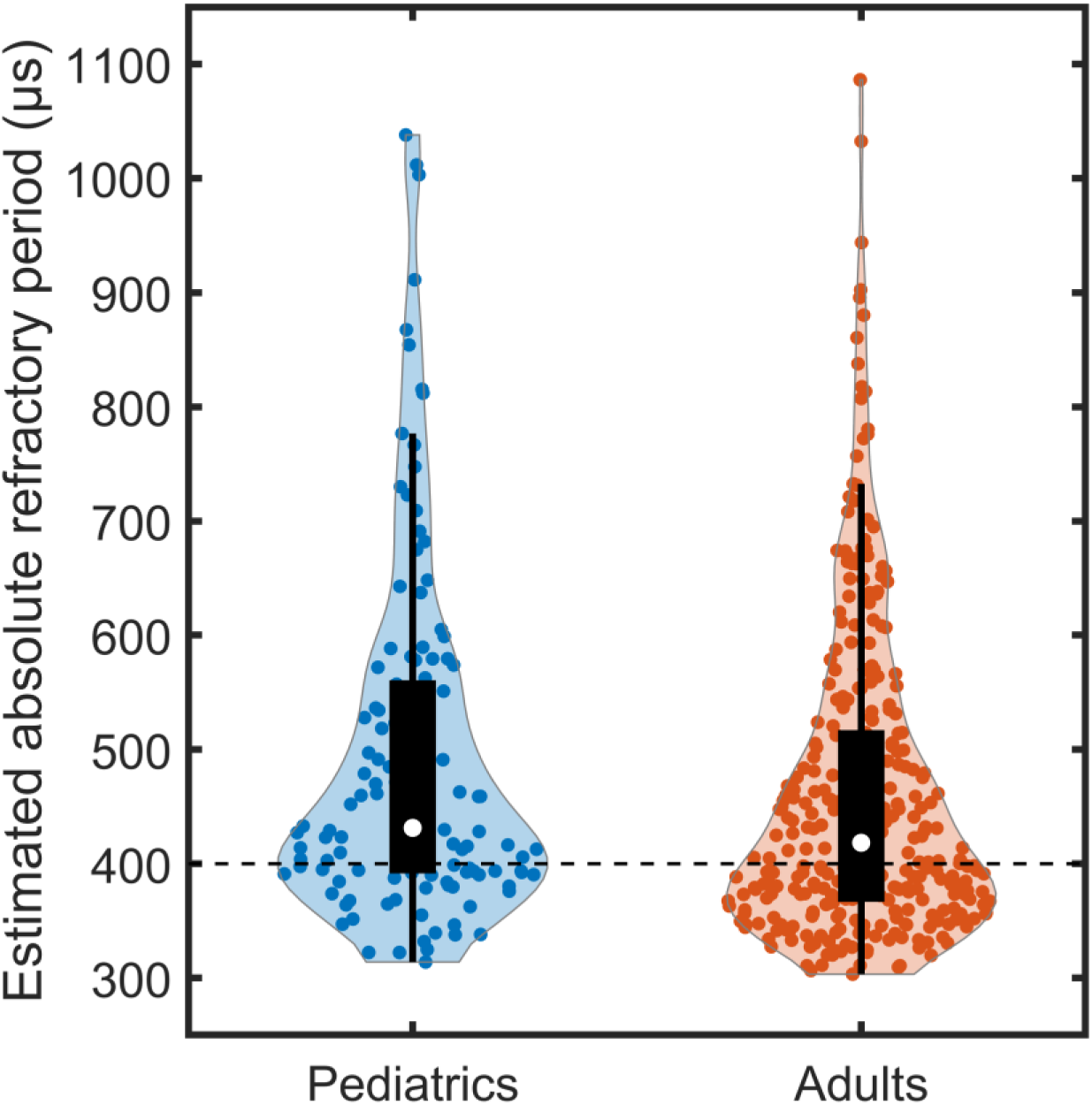
Violin plots of absolute refractory periods estimated from electrically evoked compound action potential refractory recovery functions measured at 127 electrode locations in 27 pediatric cochlear implant (CI) users (blue circles) and 346 electrode locations in 53 adult CI users (red circles). The white circle represents the median value. The black box represents the interquartile range (IQR), and the vertical black lines extend to the value that is the furthest from the median while still being within 1.5*IQR from the lower or upper quartile. The dashed horizontal line at 400 µs illustrates the default masker-probe interval used in the forward masking technique for single-pulse stimulation.

To date, a viable alternative to traditional artifact rejection techniques has not been identified. As a step toward addressing this issue, we recently developed a new method for removing artifacts from eCAP recordings measured for cathodic-leading stimulation.

## METHODS

### Study Participants

The development and validation of the new method for removing artifacts from eCAP recordings was performed in a subset of the 80 pediatric and adult CI users in whom estimates of the ARP were obtained at multiple electrode locations (see subsection Absolute Refractory Periods at High Stimulation Levels above). Specifically, this subset of CI users included 17 pediatric and adult CI users (8 Female, 9 Male) ranging in age from 16.9 to 84.0 years (mean: 53.5 years, SD: 22.1 years). Participants A3, A5, and P2 were implanted bilaterally, and each ear was tested separately in this study. Additional eCAPs were measured at the two electrode locations with the largest difference in the estimated ARP in each of the 20 ears tested. Across all ears tested, there were 20 electrodes tested with an estimated ARP less than 400 µs and 20 electrodes tested with an estimated ARP greater than 400 µs. Demographic information for each of the 17 study participants, along with the estimated ARP obtained at each of the electrodes tested in this study, are provided in Table 1. Written informed consent and/or verbal assent was obtained from all study participants and/or their legal guardians at the time of data collection. The study was approved by the Biomedical Institutional Review Board (IRB) at The Ohio State University (IRB study #: 2017H0131).

**TABLE 1.** Demographic information of all study participants. CI24RE (CA), Freedom Contour Advance electrode array; SHL, sudden hearing loss; AN, acoustic neuroma; EVA, enlarged vestibular aqueduct; ARP, absolute refractory period.

| Participant ID | Sex | Ear tested | Age (years) | Internal device and electrode array | Etiology of hearing loss | Electrodes tested | Estimated ARP (μs) |
| --- | --- | --- | --- | --- | --- | --- | --- |
| A1 | M | L | 60s | CI512 | SHL | 3, 9 | 392, 392 |
| A2 | M | L | 60s | CI512 | Meniere's | 15, 18 | 378, 380 |
| A3 | F | L | 60s | CI24RE (CA) | Hereditary | 3, 21 | 330, 377 |
| A3 | F | R | 60s | CI24RE (CA) | Hereditary | 9, 15 | 353, 309 |
| A4 | F | L | 30s | CI24RE (CA) | Trauma | 3, 9 | 414, 464 |
| A5 | F | L | 50s | CI532 | Unknown | 4, 15 | 656, 349 |
| A5 | F | R | 50s | CI24RE (CA) | Unknown | 9, 21 | 485, 513 |
| A6 | F | R | 50s | CI24RE (CA) | Hereditary | 3, 21 | 387, 454 |
| A7 | M | L | 60s | CI632 | Unknown | 3, 9 | 579, 429 |
| A8 | M | L | 60s | CI532 | AN | 9, 15 | 501, 375 |
| A9 | F | R | 80s | CI532 | Hereditary | 3, 7 | 366, 359 |
| A10 | F | L | 30s | CI532 | Unknown | 3, 9 | 554, 903 |
| A11 | F | R | 50s | CI532 | Hereditary | 12, 15 | 357, 358 |
| A12 | M | R | 80s | CI632 | Unknown | 3, 9 | 441, 447 |
| A13 | F | L | 50s | CI632 | Unknown | 3, 15 | 944, 713 |
| A14 | M | L | 70s | CI632 | Unknown | 15, 21 | 389, 358 |
| P1 | M | R | 10s | CI24RE (CA) | Connexin | 4, 12 | 463, 380 |
| P2 | M | L | 10s | CI24RE (CA) | Usher | 14, 21 | 342, 350 |
| P2 | M | R | 10s | CI24RE (CA) | Usher | 2, 9 | 413, 467 |
| P3 | M | L | 10s | CI24RE (CA) | EVA | 12, 21 | 1321, 403 |

### eCAP Measurements

All eCAPs were obtained following the same procedures as those used in our previous studies [e.g., 8, 9, 40] and using the Custom Sound EP (v. 5.1 or 6.0) software interface (Cochlear Ltd, Sydney, NSW, Australia). The stimulus was one cathodic-leading, biphasic pulse with an interphase gap of 7 µs and a pulse phase duration of 25 µs/phase. The stimulus was presented at the participants’ C level for all electrodes tested and repeated four times for calculating test-retest reliability. All stimuli were presented in a monopolar-coupled stimulation mode to individual CI electrodes via an N6 sound processor connected to a programming pod. For all eCAP measurements, the recording window was set to 64 samples (3,123 µs), the longest recording window allowed in Custom Sound EP. Additional recording parameters were a 122-µs recording delay, an amplifier gain of 50 dB, a sampling rate of 20,492 Hz, and 50 sweeps per averaged eCAP response.

### Hyperbola-fitting Artifact Subtraction Method

The hyperbola-fitting artifact subtraction method (HyperFit) was developed to address the limitations of FwdMsk. This method is based on the discovery that the waveform of the combined stimulation and recording artifacts is best characterized as a hyperbola for stimulation at a single electrode location (e.g., co-located masker and probe). Therefore, we first discuss the characterization of the artifact template as a hyperbola. We then provide a conceptual overview of the new method and detail how the method was implemented in this study.

#### Artifact Characterization for Co-located Masker and Probe

We investigated the morphology of the combined stimulation and recording artifacts (i.e., artifact template) by analyzing the 100 recordings obtained using FwdMsk at electrode locations where the estimated ARP at C level was greater than 400 µs. Additionally, the masker offset was +10 CL for each of these recordings. Therefore, the artifact template obtained via FwdMsk for each of these recordings should be free of neural responses. For each recording, the artifact template was calculated by adding and subtracting the traces obtained using FwdMsk (see Figure 1). Representative artifact templates obtained at seven electrode locations in five CI users are shown in Figure 5A.

**Figure 5.**
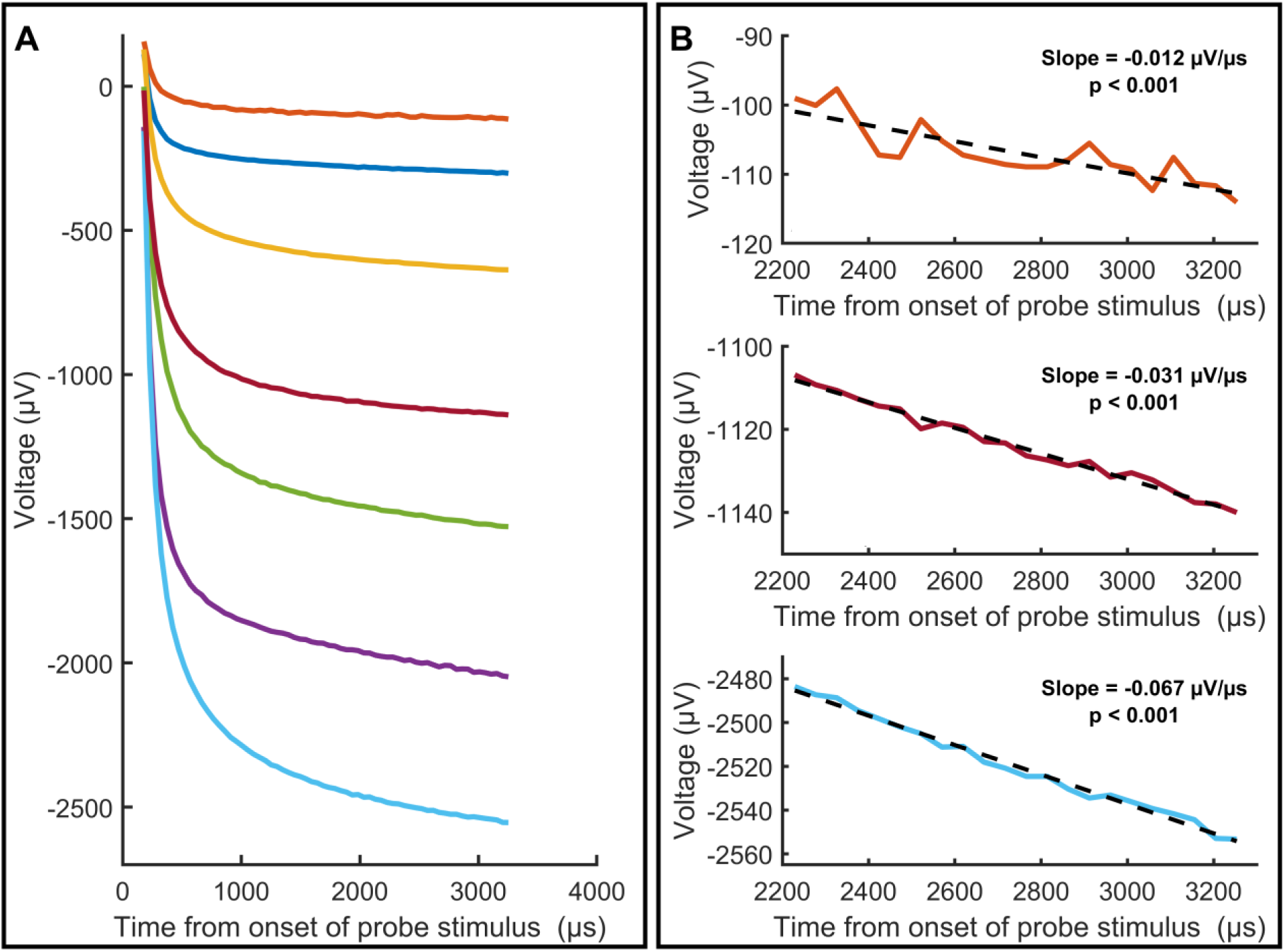
Representative artifact templates derived from recordings of the electrically evoked compound action potential (eCAP) using the classic two-pulse forward masking artifact rejection technique. Panel A: Artifact templates (colored traces) derived from eCAP recordings obtained at seven electrode locations in five cochlear implant users. Panel B: The results of linear regression using only the section of artifact template occurring after 2200 µs for three of the artifact templates shown in Panel A.

As can be seen in the figure, the artifact templates decrease monotonically with a rapid decay at the beginning of the recording window and then gradually transition to a line with a negative slope at the end of the recording window for all stimulation levels. To verify that the final section of the artifact templates reached a linear asymptote, linear regression was performed on the section of the artifact template occurring after 2200 µs. The results of linear regression for sections of three of the artifact templates shown in Figure 5A are provided in Figure 5B. Each subpanel shows a significant negative slope, and the residuals appear to be normally distributed without any systematic bias. Results of linear regression revealed that the slope was negative and significant (p ≤ 0.008), and the residuals were normally distributed as verified by the Anderson-Darling test (p ≥ 0. 538) for all 100 eCAP recordings after correcting for multiple comparisons using the False Discovery Rate [42]. Therefore, these results confirmed that the artifact template reached a slanted asymptote for all recordings.

These features observed in the artifact template (i.e., rapid initial decay and slanted asymptote) can be well described by a hyperbola. A hyperbola is a smooth curve that is described by the rational function 𝑦 = 𝑎𝑥 + 𝑏 + 𝑐(𝑥 + 𝑑)^−1^. This function has a vertical asymptote at 𝑥 = −𝑑 and a slanted asymptote at 𝑦 = 𝑎𝑥 + 𝑏. Therefore, parameter 𝑑 of the function corresponds to the vertical asymptote that bounds the rapid initial decay, while parameters 𝑎 and 𝑏 correspond to the slope and the vertical offset of the line at the latter end of the recording, respectively. Parameter 𝑐 reflects the speed of the transition between those two portions of the recording, where a larger value of 𝑐 corresponds to a slower transition. A summary of the parameter values obtained when fitting a hyperbola each of the 100 artifact templates is provided in Table 2.

**Table 2.** Summary of the parameter values obtained when fitting a hyperbola to artifact templates.

| | a ($\mu\text{V}/\mu\text{s}$) | b ($\mu\text{V}$) | c ($\mu\text{V}\cdot\mu\text{s}$) | d ($\mu\text{s}$) |
| --- | --- | --- | --- | --- |
| Minimum | -0.051 | -2,457.6 | 18,663 | -119.4 |
| Maximum | -0.002 | -94.3 | 205,970 | -58.1 |
| Mean | -0.019 | -757.8 | 67,631 | -96.1 |
| Standard deviation | 0.016 | 618.4 | 50,992 | 18.5 |

To verify that a hyperbola best describes the artifact template, we compared the goodness of fit (i.e., R^2^) obtained with a hyperbola for all 100 recordings to the goodness of fit obtained with two other functions that have been proposed to represent the artifact template: a two-component exponential function 𝑦 = 𝑎𝑒^𝑏𝑥^ + 𝑐𝑒^𝑑𝑥^ + 𝑒 [24] and a combined exponential and linear function 𝑦 = 𝑎𝑒^𝑏𝑥^ + 𝑐𝑥 + 𝑑 [43]. The R^2^s were not normally distributed for any of the three functions, so we report medians and interquartile ranges instead of means and standard deviations. A Friedman test was also conducted to determine whether the R^2^ differed between the three function fittings. As expected, the R^2^ was higher for the hyperbola than for the other two functions (Hyperbola: median = 0.999, IQR = 0.001; Two-component exponential function: median = 0.671, IQR = 0.578; Combined exponential and linear function: median = 0.991, IQR = 0.004). The result of the Friedman test showed a significant difference between the three fitting functions (χ^2^_2,198_ = 123.5, p < 0.001). All post hoc comparisons were also significant (p < 0.001 for all comparisons). Therefore, these results confirm that a hyperbola characterizes the artifact template better than the other two functions.

#### Conceptual Overview of the Hyperbola-fitting Artifact Subtraction Method

The method is illustrated in Figure 6 and entails creating an artifact template for individual recordings by fitting a hyperbola to the probe-only recording trace with greater fitting-weight given to data points in the sections of the recording in which little or no neural response is present. Specifically, greater fitting-weight is given to the data points in the recording in which the time from the onset of the probe stimulus is less than 200 µs or greater than 2200 µs. These time periods are chosen because the neural response is assumed to be very small and/or not present in these time periods. Even if there is some neural response in recording within the first 200 µs after the onset of the probe stimulus, the stimulation artifact is much larger than the neural response. A cutoff time of 2200 µs is chosen as a conservative estimate of the time point when the eCAP response has ended, which has been estimated up to 1300 µs [44, 45]. Moreover, this portion of the recording window is characterized by a line with a negative slope (see previous subsection). Therefore, we assume that the neural response should be very small or not present in that portion of the recording. After fitting the hyperbola, the hyperbola (i.e., artifact template) is subtracted from the probe-only recording to give the artifact-free eCAP waveform.

**Figure 6.**
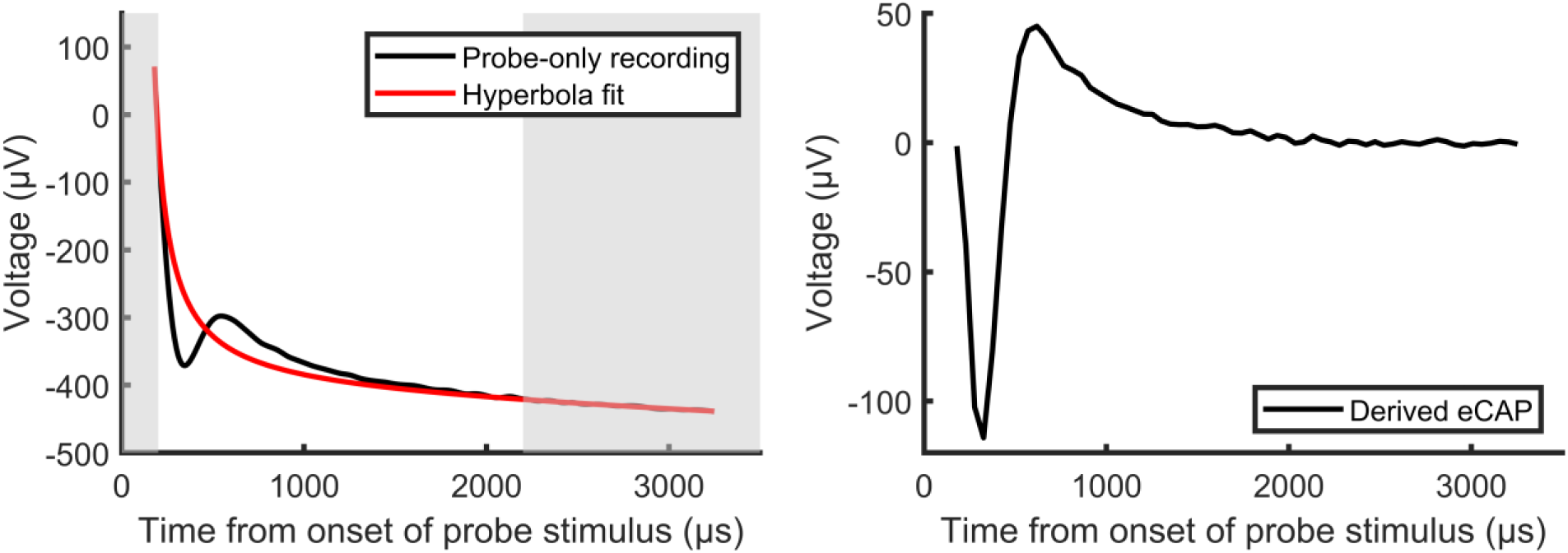
Illustration of the hyperbola-fitting artifact subtraction technique. The electrically evoked compound action potential (eCAP) waveform (black line, right panel) is derived by subtracting the hyperbola (red line, left panel) that is fit to the probe-only recording (black line, left panel) with greater fitting-weight given to data points within the time periods in which little or no neural response is present (lightly shaded regions, left panel).

#### Implementation of the Hyperbola-fitting Artifact Subtraction Method

The new method was implemented in this study in a series of three steps using MATLAB (v. 2021b) software (MathWorks Inc.).

##### Step 1: Re-sample the probe-only recording at 102,460 Hz

The default sampling frequency used in Custom Sound EP is 20,492 Hz, which corresponds to a sampling period of 48.8 µs. Therefore, for stimulation and recording parameters used in this study (i.e., pulse phase duration = 25 µs/phase, interphase gap = 7 µs, recording delay = 122 µs), only one sample was obtained within the first 200 µs after the stimulus onset. Specifically, the first sample occurred at 179 µs and the second sample occurred at 227.8 µs after the stimulus onset. Therefore, the probe-only recording was resampled at 102,460 Hz (i.e., 5x the original sampling rate) using spline interpolation to obtain a sampling resolution of 9.8 µs, which provided three data points within the first 200 µs after the stimulus onset. The resampling was done using the ‘spline’ method of the ‘interp1’ MATLAB function which uses cubic spline interpolation.

##### Step 2: Fit the hyperbola to the re-sampled waveform with custom weighting values

In typical function-fitting, each data point is given equal weight in the least-squares error minimization. However, due to the presence of both artifact and neural response in the probe-only recordings, it was necessary to use custom weighting values to emphasis the fitting to the portions of the recording in which little or no neural response is present. Specifically, the data points in the recording in which the time from stimulus onset is less than 200 µs or greater than 2200 µs were given the standard weight of 1, while all other data points were given a weight of 0.015. This weighting emphasized the fitting of the vertical asymptote (i.e., fitting-parameter 𝑑) and the slanted asymptote (i.e., fitting-parameters 𝑎 and 𝑏) while also allowing the remaining data points, even if they contain substantial neural response, to guide the transition between the asymptotes (i.e., fitting-parameter 𝑐). The function fitting was done using the ‘fit’ function of MATLAB’s Curve Fitting Toolbox. Other than the custom weighting values, all other fitting options were the default options.

##### Step 3: Subtract the hyperbola from the original probe-only recording

Importantly, this subtraction is performed only at the original sampling times. Therefore, this method does not modify the probe-only recording when deriving the eCAP waveform. Rather, the up-sampling simply creates more data points for the fitting process.

### Comparison between Forward Masking and New Method

In theory, eCAP amplitudes obtained using FwdMsk and HyperFit should be similar in cases where the masker offset is sufficiently large, the estimated ARP is greater than the MPI, and the masker and probe pulses are presented at the same electrode location. In contrast, a difference in eCAP amplitudes obtained using the two methods would be expected if the estimated ARP were less than the MPI, with greater differences in eCAP amplitudes observed for greater differences between the estimated ARP and the MPI. We tested these theoretical expectations by comparing eCAP amplitudes obtained using FwdMsk and HyperFit at each of the 40 electrodes tested.

### Statistical Analyses

The theoretical expectations were assessed using Linear Mixed-effects Models (LMMs). Specifically, the effect of the artifact removal method on the eCAP amplitude was assessed with a LMM where the eCAP amplitude was the outcome variable, the artifact removal method was the fixed effect, and participant, electrode location and test ear (i.e., Left/Right) were random effects. One LMM was used with the data for which the estimated ARP was less than 400 µs and one used with the data for which the estimated ARP was greater than 400 µs according to the theoretical expectations. A third LMM assessed the effect of the estimated ARP on the difference in the eCAP amplitude obtained using the two artifact removal methods when the estimated ARP was less than 400 µs. For this LMM, the difference in the eCAP amplitude obtained using the two methods (FwdMsk – HyperFit) was the outcome variable, the estimated ARP was the fixed effect, and participant, electrode location and test ear were random effects.

The test-retest reliability of the fitting-parameters and the eCAP amplitudes obtained using HyperFit for repeated measurements of the same stimulus were evaluated with the intraclass correlation coefficient (ICC). ICC(2,1) was chosen as the metric of interest because it quantifies the level of agreement across trials [46]. All statistical analyses for this study were performed using MATLAB (v. 2021b) software (MathWorks Inc.).

## RESULTS AND DISCUSSION

Representative eCAP waveforms derived using FwdMsk and HyperFit are shown in the bottom panels of Figure 7 and Figure 8 from recordings at three electrode locations at which the estimated ARP was greater than 400 µs and less than 400 µs, respectively. The probe-only recordings and the artifact templates from which the eCAP waveforms were derived are shown in the top panels of each figure.

**Figure 7.**
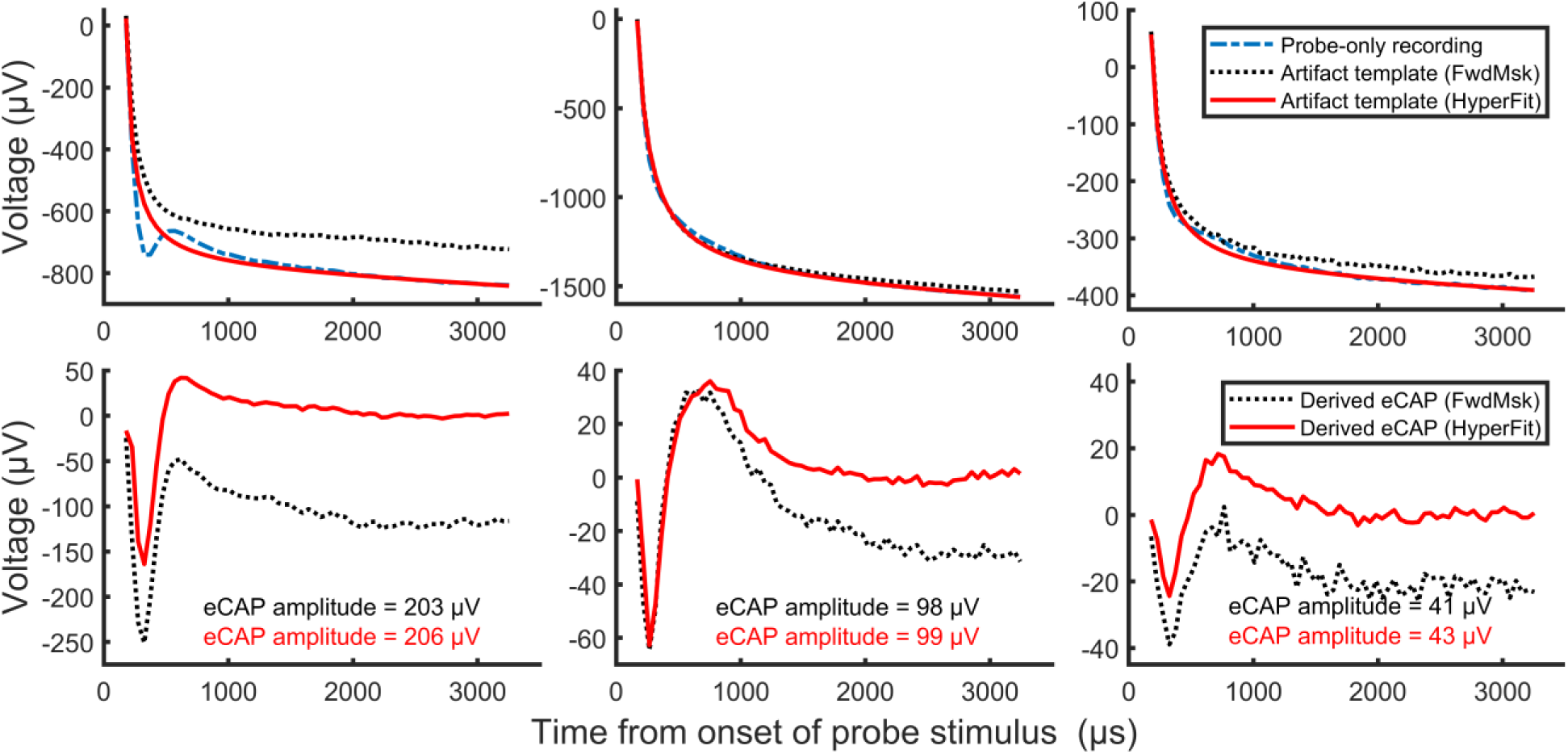
Representative probe-only recordings and artifact templates (top panels), along with electrically evoked compound action potential (eCAP) waveforms (bottom panels), obtained using the classic two-pulse forward masking artifact rejection technique (FwdMsk) and the new hyperbola-fitting artifact subtraction technique (HyperFit) at three electrode locations at which the estimated absolute refractory period (i.e., t0) was greater than 400 µs.

**Figure 8.**
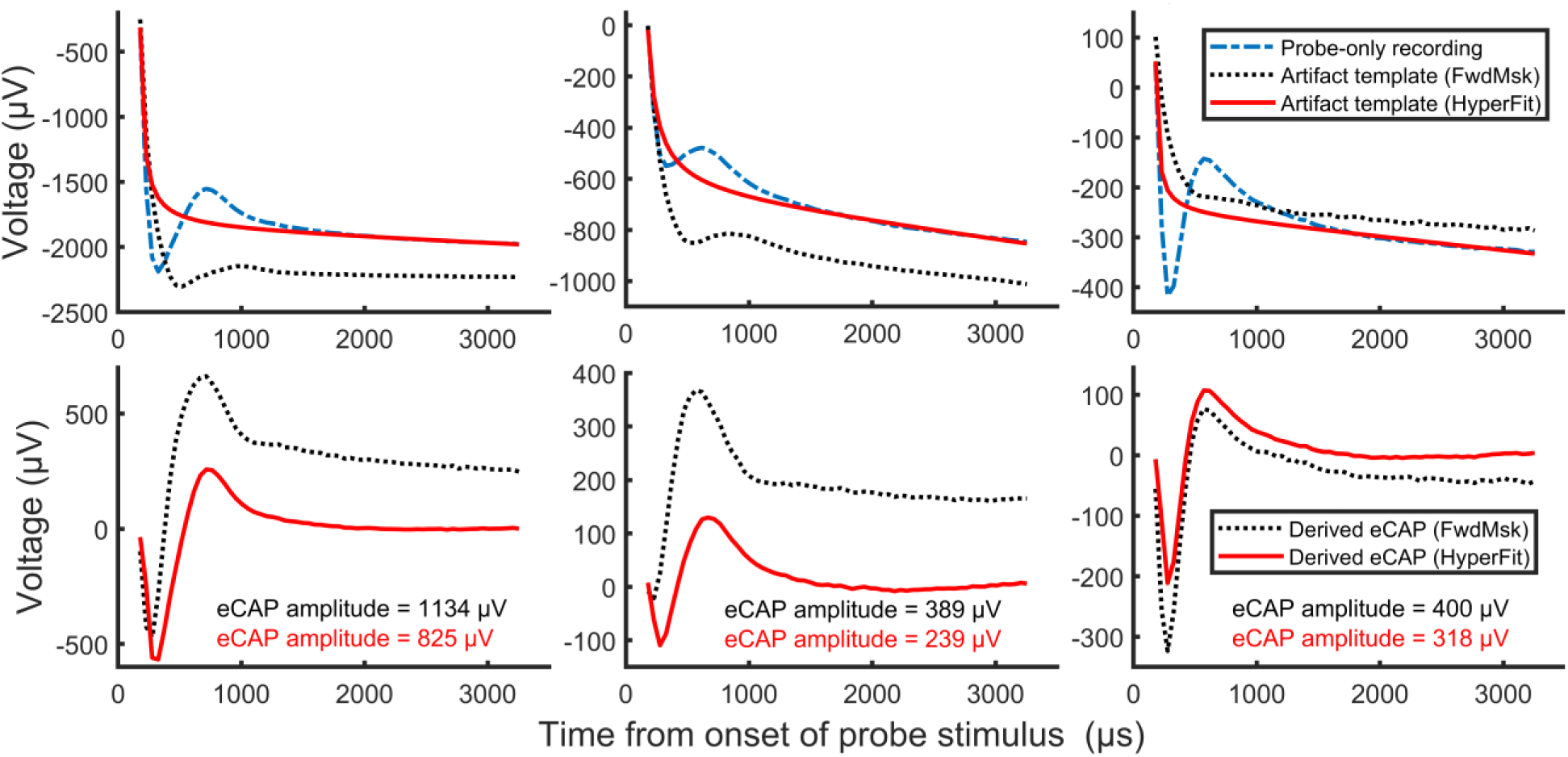
Representative probe-only recordings and artifact templates (top panels), along with electrically evoked compound action potential (eCAP) waveforms (bottom panels), obtained using the classic two-pulse forward masking artifact rejection technique (FwdMsk) and the new hyperbola-fitting artifact subtraction technique (HyperFit) at three electrode locations at which the estimated absolute refractory period (i.e., t0) was less than 400 µs.

As observed in Figure 7, the eCAP waveforms derived using FwdMsk and HyperFit were comparable when the estimated ARP was greater than 400 µs. Most importantly, the difference in eCAP amplitudes obtained using the two methods was less than 5 µV, which is within the noise floor of these devices [47]. In contrast, there were large differences in eCAP amplitudes obtained using these two methods when the estimated ARP was smaller than 400 µs, as observed in Figure 8. The primary reason for the larger eCAP amplitudes when using FwdMsk was the phase difference/shift between the neural response to the probe pulse in the masker + probe stimulation condition and the neural response included in the probe-only recording. Therefore, the neural response was present in the artifact template derived using FwdMsk, which altered the morphology of the derived eCAP waveform.

One difference in the eCAP waveforms obtained using the FwdMsk and HyperFit observed in Figure 7 and Figure 8 is the difference in plateau voltage of the eCAP waveform (i.e., vertical offset). The characteristics of the operational amplifiers included in the CI contributes, at least partially, to this vertical offset as well as the variability in the observed offset across study participants and test electrodes. Specifically, the telemetry circuitry includes an auto-zero amplifier which sets the zero/reference point shortly before the first voltage sample is acquired in each measurement trace. All subsequent samples of that trace are measured relative to that zero/reference point. Since each measurement trace has its own reference point, the vertical offset between traces is not consistent. The eCAP waveform obtained using FwdMsk is the result of subtracting four recording traces, while only one recording trace is used in the HyperFit method. This methodological difference results in a vertical offset between the derived waveforms. Another potential factor that might have contributed to the vertical offset is the voltage difference between the resting physiological voltage before the eCAP response and the physiological voltage after the eCAP response measured at the end of the probe-only recording. Any voltage difference would have been captured in the artifact template obtained using HyperFit but might have not been captured in the artifact template obtained using FwdMsk. However, there is no scientific evidence so far supporting the existence of a difference in physiological voltage before and after the eCAP response. In general, the characteristics of a voltage offset may have scientific or clinical value, but this remains unknown. In contrast, the eCAP amplitude has been used frequently in scientific and clinical studies. Therefore, we use the eCAP amplitude as a metric for comparing FwdMsk and HyperFit in this study.

The difference in the eCAP amplitude obtained using FwdMsk and HyperFit as a function of the estimated ARP is shown in Figure 9 for all 40 electrode locations tested. As can be observed in the figure, the difference in the eCAP amplitude was near zero for all electrode locations at which the estimated ARP was greater than 400 µs (i.e., the MPI of the stimulus). In contrast, for electrode locations at which the estimated ARP was less than 400 µs, the difference in the eCAP amplitude increased with decreasing ARP. These observations were confirmed by the results of statistical analyses. Specifically, there was not an effect of the artifact removal method on the eCAP amplitude when the estimated ARP was greater than 400 µs (F_1,38_ = 0.19, p = 0.666). In contrast, there was a significant effect of the artifact removal method on the eCAP amplitude when the estimated ARP was less than 400 µs (F_1,38_ = 13.67, p < 0.001). Finally, there was a significant effect of the estimated ARP on the difference in the eCAP amplitude when the estimated ARP was less than 400 µs (F_1,18_ = 68.19, p < 0.001). These experimental results match the results expected based on theory.

**Figure 9.**
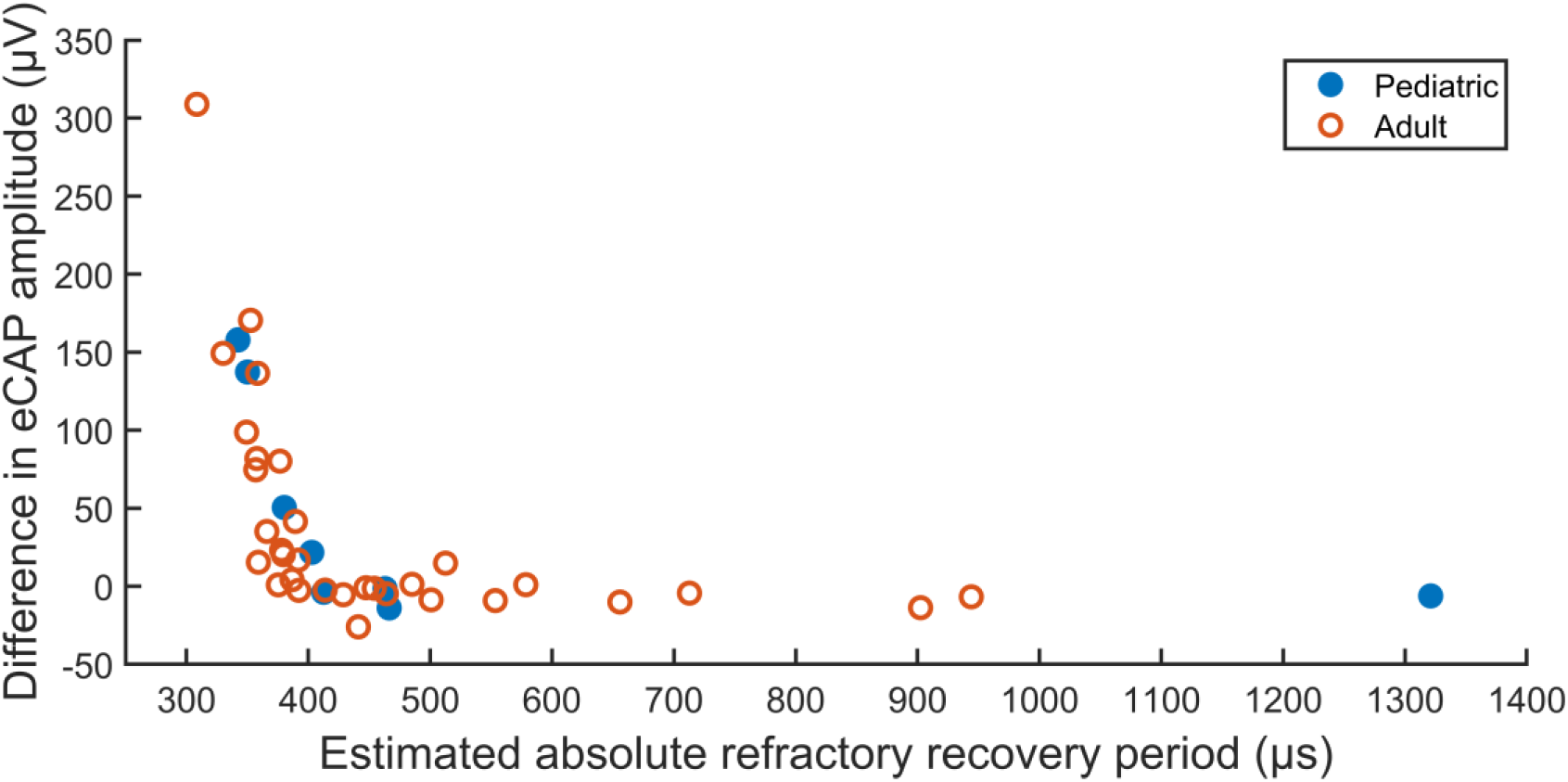
The difference in electrically evoked compound action potential (eCAP) amplitude obtained using the classic two-pulse forward masking artifact rejection technique and the new hyperbola-fitting artifact subtraction technique at two electrode locations per test ear in four ears of three pediatric cochlear implant (CI) users (blue filled circles) and 16 ears of 14 adult CI users (red unfilled circles) as a function of the estimated absolute refractory recovery period.

### Test-retest Reliability

Estimates of intraclass correlation coefficients, along with the 95% confidence intervals, for fitting-parameters and eCAP amplitudes obtained using HyperFit are provided in Table 3. Clearly, there is excellent test-retest reliability for all fitting-parameters and the eCAP amplitude obtained using HyperFit.

**Table 3.** Intraclass correlation coefficients for fitting-parameters and electrically evoked compound action potential (eCAP) amplitudes obtained using the new method.

|  | Fitting parameters |  |  |  | eCAP amplitude |
| --- | --- | --- | --- | --- | --- |
|  | a | b | c | d |  |
| Estimate | 0.987 | 0.998 | 0.999 | 0.991 | 0.995 |
| Confidence Interval | [0.975, 0.993] | [0.997, 0.999] | [0.998, 1.000] | [0.985, 0.995] | [0.990, 0.998] |

## CONCLUSIONS

The underlying assumption of the classic two-pulse forward masking is invalid in up to 45% of cases at high stimulation levels. Additionally, the eCAP amplitude obtained using the forward masking technique is highly affected by varying the masker offset or the masker-probe interval. Therefore, it is important to verify appropriate stimulation settings (i.e., masker level and masker-probe interval) in any study that uses the forward masking technique to calculate eCAP amplitudes. This is especially important in studies that use the eCAP amplitude as a parameter for predicting neural health or as a correlate to results of auditory perception. For cases in which the assumptions of forward masking are met (i.e., +10 CL masker offset and ARP > MPI), the eCAP amplitudes obtained using forward masking are comparable to the eCAP amplitudes obtained using the new method. Additionally, the eCAP amplitude calculated using the new method is consistent across repeated measurements. Therefore, the new method presented in this report is a viable alternative to the forward masking technique for obtaining artifact-free eCAPs in cathodic-leading, single-pulse stimulation. Moreover, it has the advantage of reduced recording time because it only requires one recording trace (vs. 4 required by the forward masking technique), and eCAPs can be recorded at higher stimulation levels because it does not require a strong masker pulse. The new method has currently only been validated for eCAPs evoked by single, cathodic-leading, biphasic pulses with fixed stimulation parameters (e.g., pulse phase duration = 25 µs/phase, inter-phase gap = 7 µs) in patients with normal inner-ear anatomy that were implanted with a Cochlear™ Nucleus® CI (Cochlear Ltd.). Research investigating the application of the new method with other stimulation parameters and testing paradigms (e.g., spread of excitation, refractory recovery, pulse-train stimulation) in various CI patient populations, along with optimization of the parameters of the new method (e.g., fitting-weights), is in process. Additionally, future studies will evaluate the relationships between eCAP metrics obtained using the new method (e.g., eCAP threshold) and behavioral measures (e.g., detection thresholds).

## Author Contributions

Jeffrey Skidmore: Conceptualization, Methodology, Investigation, Writing - Original draft preparation, Visualization. Yi Yuan: Investigation, Writing – Review and editing. Shuman He: Funding acquisition, Conceptualization, Supervision, Writing - Review and editing.

## Data Availability

All data produced in the present study are available upon reasonable request to the authors

## Acknowledgments

The authors thank Ryan Melman (Senior Research Engineer, Cochlear Ltd.) for an insightful discussion about operational amplifiers and artifact removal methods

## REFERENCES

1. Brown CJ, Abbas PJ, and Gantz B. Electrically evoked whole-nerve action potentials: Data from human cochlear implant users. The Journal of the Acoustical Society of America. 1990;88(3):1385–91.

2. He S, Teagle HFB, and Buchman CA. The Electrically Evoked Compound Action Potential: From Laboratory to Clinic. Frontiers in Neuroscience. 2017;11:339.

3. Brochier T, Guérit F, Deeks JM, Garcia C, Bance M, and Carlyon RP. Evaluating and comparing behavioural and electrophysiological estimates of neural health in cochlear implant users. Journal of the Association for Research in Otolaryngology. 2021;22(1):67–80.

4. Jahn KN, and Arenberg JG. Electrophysiological Estimates of the Electrode-Neuron Interface Differ Between Younger and Older Listeners With Cochlear Implants. Ear and Hearing. 2020;41(4):948–60.

5. Jahn KN, and Arenberg JG. Identifying cochlear implant channels with relatively poor electrode-neuron interfaces using the electrically evoked compound action potential. Ear and Hearing. 2020;41(4):961–73.

6. Schvartz-Leyzac KC, Holden TA, Zwolan TA, Arts HA, Firszt JB, Buswinka CJ, et al. Effects of Electrode Location on Estimates of Neural Health in Humans with Cochlear Implants. Journal of the Association for Research in Otolaryngology. 2020;21(3):259–75.

7. Garcia C, Goehring T, Cosentino S, Turner RE, Deeks JM, Brochier T, et al. The Panoramic ECAP Method: Estimating Patient-Specific Patterns of Current Spread and Neural Health in Cochlear Implant Users. Journal of the Association for Research in Otolaryngology. 2021;22(5):567–89.

8. Skidmore J, Carter BL, Riggs WJ, and He S. The effect of advanced age on the electrode-neuron interface in cochlear implant users. Ear and Hearing. 2022;43(4):1300–15.

9. Skidmore J, Xu L, Chao X, Riggs WJ, Pellittieri A, Vaughan C, et al. Prediction of the functional status of the cochlear nerve in individual cochlear implant users using machine learning and electrophysiological measures. Ear and Hearing. 2021;42(1):180–92.

10. Schvartz-Leyzac KC, Colesa DJ, Buswinka CJ, Swiderski DL, Raphael Y, and Pfingst BE. Changes over time in the electrically evoked compound action potential (ECAP) interphase gap (IPG) effect following cochlear implantation in Guinea pigs. Hearing Research. 2019;383:107809.

11. Ramekers D, Benav H, Klis SFL, and Versnel H. Changes in the electrically evoked compound action potential over time after implantation and subsequent deafening in guinea pigs. Journal of the Association for Research in Otolaryngology. 2022.

12. Schvartz-Leyzac KC, Zwolan TA, and Pfingst BE. Using the electrically-evoked compound action potential (ECAP) interphase gap effect to select electrode stimulation sites in cochlear implant users. Hearing Research. 2021;406.

13. Müller-Deile J, Neben N, Dillier N, Büchner A, Mewes A, Junge F, et al. Comparisons of electrophysiological and psychophysical fitting methods for cochlear implants. International Journal of Audiology. 2023;62(2):118–28.

14. Chao X, Wang R, Luo J, Wang H, Fan Z, and Xu L. Relationship between electrically evoked compound action potential thresholds and behavioral T-levels in implanted children with cochlear nerve deficiency. Scientific Reports. 2023;13(1):4309.

15. Dong Y, Briaire JJ, Stronks HC, and Frijns JHM. Speech Perception Performance in Cochlear Implant Recipients Correlates to the Number and Synchrony of Excited Auditory Nerve Fibers Derived From Electrically Evoked Compound Action Potentials. Ear and Hearing. 2023;44(2):276–86.

16. Skidmore J, Oleson JJ, Yuan Y, and He S. The Relationship Between Cochlear Implant Speech Perception Outcomes and Electrophysiological Measures of the Electrically Evoked Compound Action Potential. Ear and Hearing. 2023;44(6):1485–97.

17. Skidmore J, Ramekers D, Colesa DJ, Schvartz-Leyzac KC, Pfingst BE, and He S. A broadly applicable method for characterizing the slope of the electrically evoked compound action potential amplitude growth function. Ear and Hearing. 2022;43(1):150–64.

18. He S, Skidmore J, Carter BL, Lemeshow S, and Sun S. Postlingually Deafened Adult Cochlear Implant Users With Prolonged Recovery From Neural Adaptation at the Level of the Auditory Nerve Tend to Have Poorer Speech Perception Performance. Ear and Hearing. 2022;43(6):1761–70.

19. Botros A, and Psarros C. Neural response telemetry reconsidered: II. the influence of neural population on the ECAP recovery function and refractoriness. Ear and Hearing. 2010;31(3):380–91.

20. Geddes LA. Historical evolution of circuit models for the electrode-electrolyte interface. Annals of Biomedical Engineering. 1997;25(1):1–14.

21. Miller CA, Abbas PJ, Rubinstein JT, Robinson BK, Matsuoka AJ, and Woodworth G. Electrically evoked compound action potentials of guinea pig and cat: Responses to monopolar, monophasic stimulation. Hearing Research. 1998;119(1-2):142–54.

22. Bahmer A, Peter O, and Baumann U. Recording and analysis of electrically evoked compound action potentials (ECAPs) with MED-EL cochlear implants and different artifact reduction strategies in Matlab. Journal of Neuroscience Methods. 2010;191(1):66–74.

23. Akhoun I, McKay CM, and El-Deredy W. Electrically evoked compound action potential artifact rejection by independent component analysis: technique validation. Hearing Research. 2013;302:60–73.

24. Spitzer P, Zierhofer C, and Hochmair E. Algorithm for multi-curve-fitting with shared parameters and a possible application in evoked compound action potential measurements. BioMedical Engineering OnLine. 2006;5:13.

25. Baudhuin JL, Hughes ML, and Goehring JL. A Comparison of Alternating Polarity and Forward Masking Artifact-Reduction Methods to Resolve the Electrically Evoked Compound Action Potential. Ear and Hearing. 2016;37(4):e247–55.

26. Miller CA, Abbas PJ, and Brown CJ. An improved method of reducing stimulus artifact in the electrically evoked whole-nerve potential. Ear and Hearing. 2000;21(4):280–90.

27. He S, Xu L, Skidmore J, Chao X, Jeng F-C, Wang R, et al. The effect of interphase gap on neural response of the electrically stimulated cochlear nerve in children with cochlear nerve deficiency and children with normal-sized cochlear nerves. Ear and Hearing. 2020;41(4):918–34.

28. He S, Xu L, Skidmore J, Chao X, Riggs WJ, Wang R, et al. Effect of increasing pulse phase duration on neural responsiveness of the electrically stimulated cochlear nerve. Ear and Hearing. 2020;41(6):1606–18.

29. Xu L, Skidmore J, Luo J, Chao X, Wang R, Wang H, et al. The effect of pulse polarity on neural response of the electrically stimulated cochlear nerve in children with cochlear nerve deficiency and children with normal-sized cochlear nerves. Ear and Hearing. 2020;41(5):1306–19.

30. Luo J, Xu L, Chao X, Wang R, Pellittieri A, Bai X, et al. The effects of GJB2 or SLC26A4 gene mutations on neural response of the electrically stimulated auditory nerve in children. Ear and Hearing. 2020;41(1):194–207.

31. Hughes ML. Electrically evoked compound action potential polarity sensitivity, refractory-recovery, and behavioral multi-pulse integration as potential indices of neural health in cochlear-implant recipients. Hearing Research. 2023;433:108764.

32. Hughes ML. Characterizing polarity sensitivity in cochlear implant recipients: Demographic effects and potential implications for estimating neural health. Journal of the Association for Research in Otolaryngology. 2022;23(2):301–18.

33. Hughes ML, Werff KRV, Brown CJ, Abbas PJ, Kelsay DMR, Teagle HFB, et al. A longitudinal study of electrode impedance, the electrically evoked compound action potential, and behavioral measures in Nucleus 24 cochlear implant users. Ear and Hearing. 2001;22(6):471–86.

34. Miller CA, Abbas PJ, and Robinson BK. Response properties of the refractory auditory nerve fiber. Journal of the Association for Research in Otolaryngology. 2001;2(3):216–32.

35. Tabibi S, Kegel A, Lai WK, Bruce IC, and Dillier N. Measuring temporal response properties of auditory nerve fibers in cochlear implant recipients. Hearing Research. 2019;380:187–96.

36. Boulet J, White M, and Bruce IC. Temporal considerations for stimulating spiral ganglion neurons with cochlear implants. Journal of the Association for Research in Otolaryngology. 2016;17(1):1–17.

37. Hey M, Müller-Deile J, Hessel H, and Killian M. Facilitation and refractoriness of the electrically evoked compound action potential. Hearing Research. 2017;355:14–22.

38. Cartee LA, Miller CA, and van den Honert C. Spiral ganglion cell site of excitation I: Comparison of scala tympani and intrameatal electrode responses. Hearing Research. 2006;215(1-2):10–21.

39. Ramekers D, Versnel H, Strahl SB, Klis SFL, and Grolman W. Recovery characteristics of the electrically stimulated auditory nerve in deafened guinea pigs: Relation to neuronal status. Hearing Research. 2015;321:12–24.

40. He S, Shahsavarani BS, McFayden TC, Wang H, Gill KE, Xu L, et al. Responsiveness of the electrically stimulated cochlear nerve in children with cochlear nerve deficiency. Ear and Hearing. 2018;39(2):238–50.

41. Morsnowski A, Charasse B, Collet L, Killian M, and Müller-Deile J. Measuring the refractoriness of the electrically stimulated auditory nerve. Audiology and Neurotology. 2006;11(6):389–402.

42. Benjamini Y, and Hochberg Y. Controlling the false discovery rate: A practical and powerful approach to multiple testing. Journal of the Royal Statistical Society: Series B (Methodological). 1995;57(1):289–300.

43. Chakravarthy K, Fitzgerald J, Will A, Trutnau K, Corey R, Dinsmoor D, et al. A Clinical Feasibility Study of Spinal Evoked Compound Action Potential Estimation Methods. Neuromodulation: Technology at the Neural Interface. 2022;25(1):75–84.

44. Botros A, van Dijk B, and Killian M. AutoNRT: an automated system that measures ECAP thresholds with the Nucleus Freedom cochlear implant via machine intelligence. Artificial Intelligence in Medicine. 2007;40(1):15–28.

45. Gartner L, Lenarz T, and Buchner A. Fine-grain recordings of the electrically evoked compound action potential amplitude growth function in cochlear implant recipients. BioMedical Engineering OnLine. 2018;17(1):140.

46. Shrout PE, and Fleiss JL. Intraclass Correlations - Uses in Assessing Rater Reliability. Psychological Bulletin. 1979;86(2):420–8.

47. Patrick JF, Busby PA, and Gibson PJ. The development of the Nucleus Freedom Cochlear implant system. Trends in Amplification. 2006;10(4):175–200.

